# Willingness to pay for improved long-term care insurance among beneficiaries or primary family caregivers in a Chinese pilot city: A contingent valuation study

**DOI:** 10.64898/2026.05.28.26354309

**Authors:** Huai-wen Cao, Xin-yang Li, Zhi-qiang Cao

**Author notes:** Corresponding author: Xin-yang Li, Zhi-qiang Cao.

## Abstract

**Background:** China’s rapidly ageing population has increased the demand for long-term care insurance (LTCI), while the sustainability of current financing arrangements remains uncertain. Understanding willingness to pay (WTP) for improved LTCI services among LTCI beneficiaries or primary family caregivers may provide empirical evidence for discussions on acceptable and sustainable contribution mechanisms.

**Methods:** We conducted a contingent valuation survey among 278 LTCI beneficiaries or primary family caregivers in Panjin City, Liaoning Province, China. An iterative bidding game with randomized starting bids was used to elicit monthly WTP for a predefined LTCI service improvement scenario. Tobit regression models with heteroskedasticity-robust standard errors were used to estimate factors associated with WTP, including household income, disability severity, satisfaction with current services, and demographic characteristics.

**Results:** The mean monthly WTP for improved LTCI services was approximately CNY 300, compared with the current average monthly premium of approximately CNY 120. The median WTP was CNY 250. Higher household income was positively associated with WTP. Compared with participants with monthly household income below CNY 5,000, those in the highest income group above CNY 30,000 reported an additional WTP of CNY 178.9. More severe disability was also associated with higher WTP, whereas greater satisfaction with current LTCI services was associated with lower WTP. These associations were generally consistent across alternative model specifications.

**Conclusions:** LTCI beneficiaries or primary family caregivers in this Chinese pilot city reported a willingness to contribute more for improved LTCI services, particularly among those with higher income, greater care needs, or lower satisfaction with current services. These findings may inform discussions on differentiated contribution arrangements and service quality improvements in LTCI financing reform. However, the results should be interpreted cautiously because the study was conducted in a single pilot city and relied on stated-preference data.

## Introduction

Long-term care insurance (LTCI) is increasingly important for ageing societies facing growing demand for assistance with activities of daily living, chronic disease management, and long-term support services. From the perspective of social insurance, LTCI can pool risks that are difficult for individuals and households to predict or finance independently. Classic economic theory emphasizes the importance of risk pooling when individuals face uncertain and potentially catastrophic health-related expenditures [1]. In the context of long-term care, social insurance may also help address undersaving for distant care needs and reduce the financial burden placed on older adults and family caregivers [2,3].

China is experiencing rapid population ageing. National statistics and international demographic projections indicate that the proportion and absolute number of older adults in China will continue to rise in the coming decades [4,5]. At the same time, the number of older adults with functional limitations or disability is expected to increase, creating growing demand for formal long-term care services [6,7]. This demographic transition has made sustainable LTCI financing an important public health, health financing, and social policy issue.

China began piloting LTCI in selected cities in 2016 to provide care security for older adults and individuals with disability [8]. Since then, pilot programmes have explored different financing mechanisms, eligibility assessment systems, reimbursement arrangements, and service delivery models [7,9]. However, China’s LTCI system remains at an exploratory stage. Financing arrangements vary across regions, and there is not yet a single national contribution model. Balancing fiscal sustainability, affordability, equity, and service quality remains a central challenge for LTCI development in China [7,10].

International experience may provide useful reference points. Japan introduced its public LTCI system in 2000 and has since developed a financing model that combines public funding, insurance premiums, standardized eligibility assessment, and periodic institutional adjustment [11,12]. Studies of Japan’s system have also shown that cost sharing, regional variation, and demographic pressures continue to shape long-term care utilization and expenditure [13,14]. These experiences are relevant to China, but direct policy transfer should be approached cautiously because the two countries differ in institutional arrangements, demographic structure, income distribution, and fiscal capacity.

Evidence on willingness to pay (WTP) can contribute to discussions on LTCI financing. WTP reflects how individuals value potential improvements in services under specified conditions and can help identify heterogeneity in preferences and payment capacity. In health economics, contingent valuation methods are widely used to elicit WTP for health services, insurance benefits, and other nonmarket goods when revealed-preference data are unavailable or incomplete [16–19]. Because LTCI service improvements are not typically traded in a conventional market, a stated-preference approach can provide useful information on how beneficiaries or family caregivers value potential changes in service coverage and quality.

Although LTCI financing has attracted increasing research attention in China, much of the existing literature has focused on macro-level policy design, institutional evaluation, or programme implementation [7,9,10,25]. Fewer studies have directly elicited individual-level WTP among current LTCI beneficiaries or primary family caregivers using stated-preference methods. Evidence from local pilot programmes may be useful because LTCI implementation in China remains regionally diverse.

This study aimed to estimate WTP for improved LTCI services among LTCI beneficiaries or primary family caregivers in Panjin, a Chinese LTCI pilot city, using a contingent valuation approach. We also examined socioeconomic, health-related, and service-related factors associated with WTP, including household income, disability severity, satisfaction with current services, and demographic characteristics. The findings are intended to provide empirical evidence for policy discussions on LTCI financing rather than to prescribe a specific premium level.

## Materials and methods

### Study setting

Panjin is a prefecture-level city in Liaoning Province, northeastern China, with a population of approximately 1.3 million. Panjin has implemented a basic LTCI pilot scheme since 2017, covering eligible urban employees and residents. The current monthly individual premium for basic LTCI coverage averages approximately CNY 120, with additional financing from government and employer contributions.

Panjin provides a relevant local setting for examining LTCI financing because it is located in northeastern China, a region facing population ageing, industrial restructuring, and outmigration of younger working-age residents. These characteristics make the city suitable for studying beneficiary-level preferences in a local LTCI pilot context. However, Panjin should not be interpreted as nationally representative of China as a whole.

### Survey instrument

The questionnaire was designed using the contingent valuation method (CVM), a standard stated-preference approach used in health economics and other fields to value nonmarket goods and services [15,16]. CVM is commonly used to elicit WTP for health care programmes, insurance benefits, and long-term care services when market prices or revealed-preference data are unavailable or insufficient [17,18]. The survey instrument included four main sections.

First, sociodemographic characteristics were collected, including age, gender, educational level, monthly household income, district of residence, disability level, number of diagnosed chronic conditions, respondent type, and preferred care model. Monthly household income was categorized as less than CNY 5,000; CNY 5,001–10,000; CNY 10,001–20,000; CNY 20,001–30,000; and more than CNY 30,000. Disability level was classified according to the local activities of daily living assessment standard as mild, moderate, or severe. Respondent type was recorded as either the LTCI beneficiary or the primary family caregiver of the beneficiary.

Second, satisfaction with current LTCI services was assessed using seven items on a 5-point Likert scale, ranging from 1, “very dissatisfied”, to 5, “very satisfied”. The items covered overall service quality, caregiver professionalism and skills, waiting time for service provision, accessibility of care facilities, communication responsiveness, and related service domains. The overall satisfaction score was calculated as the average of the seven items. Internal consistency was acceptable, with Cronbach’s alpha equal to 0.82.

Third, WTP was elicited using a contingent valuation scenario and iterative bidding game. Participants first reported their current monthly LTCI premium contribution. They were then presented with a predefined LTCI service improvement scenario. The scenario described improvements to the LTCI service package, including higher caregiver-to-recipient staffing ratios, expanded rehabilitation and preventive health services, reduced waiting times for service initiation, improved caregiver training and certification standards, and better physical conditions at care facilities.

To reduce hypothetical bias, a brief “cheap talk” script was presented before the bidding process. This approach has been proposed as a way to reduce hypothetical bias in contingent valuation surveys [19]. Participants were asked to consider their actual household budget constraints, competing expenditures, and income limitations when stating the maximum additional amount they would be willing to pay for the described service improvements.

Participants were randomly assigned in a 1:1:1 ratio to one of three initial bid values: CNY 50, CNY 100, or CNY 150 per month above their current premium. Depending on whether the participant accepted or rejected the initial bid, the amount was adjusted upward or downward in increments of CNY 50. The bidding process ended when two directional switches occurred, when the bid interval narrowed to CNY 50 or less, or when the prespecified lower or upper bound of CNY 0 or CNY 1,000 per month was reached. The final maximum WTP was recorded as the highest accepted bid value or the midpoint of the terminal bid interval.

Participants who reported zero WTP were asked a follow-up question to distinguish genuine zero WTP from protest responses. Genuine zero responses included inability to pay because of household budget constraints or no perceived need for additional LTCI services. Protest responses included objections to the payment mechanism or beliefs that the government should fully finance LTCI through taxation. Because protest responses can bias WTP estimates if treated as genuine zero valuations, they were identified using follow-up questions [20]. No protest-zero responses were included in the final analytical sample. Genuine zero responses were retained as valid left-censored observations.

Fourth, policy awareness was assessed using items on participants’ self-reported understanding of current LTCI policies, including reimbursement rates, coverage scope, eligibility criteria, information sources, and prior exposure to LTCI public education or community outreach programmes.

The complete questionnaire was pretested with 30 participants drawn from the same LTCI registry population. Pretest feedback was used to refine item wording, validate skip logic, and ensure that the iterative bidding game could be administered consistently in both online and face-to-face survey modes.

### Sampling and data collection

A multistage stratified random sampling design was used. The sampling frame was the Panjin Municipal LTCI beneficiary registry. The registry was stratified by district of residence and disability level. District strata included Shuangtaizi, Xinglongtai, Dawa, and Panshan. Disability strata included mild, moderate, and severe disability. Within each stratum, simple random samples were drawn proportional to stratum size.

Eligible participants were LTCI beneficiaries or primary family caregivers of enrolled beneficiaries, aged 18 years or older, and able to provide informed consent. The term “participants” is used throughout the manuscript to refer to LTCI beneficiaries or primary family caregivers who completed the survey.

Data collection used a mixed-mode approach. Online self-administered surveys were distributed through secure individualized survey links with embedded eligibility screening questions and attention-check items. Face-to-face structured interviews were conducted by trained enumerators in community health centres and care facilities. Identical question wording, response options, skip patterns, and range checks were implemented across both survey modes. Random telephone back checks were performed on 10% of face-to-face interviews to monitor data quality.

Data were collected between 1 June 2024 and 31 August 2024. A total of 350 questionnaires were distributed. After excluding incomplete responses, straight-lining patterns on satisfaction scales, failed attention-check items, duplicate or ineligible responses, and other invalid questionnaires, 278 valid questionnaires were retained for the final analysis, yielding an effective response rate of 79.4%. The final analytical sample included 267 participants with positive WTP and 11 participants with genuine zero WTP. No protest-zero responses were included in the final analytical sample.

### Variables

The dependent variable was participants’ stated maximum monthly WTP for improved LTCI services, measured in Chinese yuan (CNY). Because WTP cannot be negative and genuine zero responses were retained, the dependent variable was left-censored at zero.

The main explanatory variables were selected based on economic theory and previous empirical studies of health-related WTP. Household income was included as a categorical variable, with monthly household income below CNY 5,000 as the reference group. Disability level was included as a categorical variable, with mild disability as the reference group. Overall satisfaction with current LTCI services was treated as a continuous variable based on the average score across seven Likert-scale items.

Additional covariates included age group, gender, educational level, urban residence, number of chronic conditions, preferred care model, self-reported policy awareness, respondent type, survey mode, and district fixed effects.

### Statistical analysis

Descriptive statistics were used to summarize participant characteristics, current LTCI premiums, satisfaction scores, and WTP distribution. Continuous variables were reported as means and standard deviations, while categorical variables were reported as frequencies and percentages.

Because WTP was left-censored at zero, a Type I Tobit regression model was used as the primary model [21]. The Tobit model accounts for the probability mass at the censoring point while modelling the continuous latent WTP distribution among positive observations. Heteroskedasticity-robust standard errors were used. Where appropriate, standard errors were clustered at the community health centre or care facility level to account for potential within-site correlation [22].

Several robustness checks were conducted. First, ordinary least squares regression was estimated using observed WTP, including genuine zero responses as zero-valued observations. Second, models using log-transformed WTP, ln(WTP + 1), were estimated to reduce the influence of right-tail outliers. Third, the Tobit model was reestimated after excluding genuine zero WTP observations. Fourth, subgroup analyses were conducted by disability level and survey mode. Fifth, models including starting-bid indicators were estimated to assess whether randomized starting bids were associated with final WTP estimates.

All statistical analyses were performed using Stata 18 (StataCorp LLC, College Station, TX, USA). Statistical significance was assessed using two-sided tests, with p < 0.05 considered statistically significant.

### Ethics statement

This study was conducted in accordance with the Declaration of Helsinki and was approved by the Institutional Review Board of Shenyang Medical Imaging Digital Technology Innovation Center.

Approval number: **LNKM20240504-01**

Approval date: **4 May 2024**

All participants provided informed consent before participating in the survey. For online surveys, informed consent was obtained electronically before participants accessed the questionnaire. For face-to-face interviews, written informed consent was obtained before the interview. Survey responses were anonymized at the point of data entry, and no personally identifiable information was retained in the analytical dataset. Data files were stored on encrypted, password-protected servers accessible only to the core research team.

## Results

### Participant characteristics

Table 1 summarizes the characteristics of the final analytical sample. The sample included 278 LTCI beneficiaries or primary family caregivers. The mean age of participants was 67.4 years (SD = 9.2), and 58.6% were female. Most participants lived in urban areas.

**Table 1.** Descriptive characteristics of the final analytical sample.

| Variable | Mean or % | SD | Min | Max |
| --- | --- | --- | --- | --- |
| Age, years | 67.4 | 9.2 | 42 | 89 |
| Female | 58.6% | — | — | — |
| Urban residence | 72.3% | — | — | — |
| Household monthly income < CNY 5,000 | 22.7% | — | — | — |
| Household monthly income CNY 5,001 – 10,000 | 35.6% | — | — | — |
| Household monthly income CNY 10,001 – 20,000 | 24.1% | — | — | — |
| Household monthly income CNY 20,001 – 30,000 | 11.5% | — | — | — |
| Household monthly income > CNY 30,000 | 6.1% | — | — | — |
| Mild disability | 62.2% | — | — | — |
| Moderate disability | 26.3% | — | — | — |
| Severe disability | 11.5% | — | — | — |
| Number of chronic conditions | 1.7 | 1.3 | 0 | 6 |
| Home-based care preference | 54.0% | — | — | — |
| Community-based care preference | 28.4% | — | — | — |
| Institutional care preference | 17.6% | — | — | — |
| Current monthly LTCI premium, CNY | 120 | 40 | 80 | 200 |
| Overall satisfaction score, 1 – 5 | 3.4 | 0.8 | 1.3 | 5.0 |
| Online survey mode | 52.5% | — | — | — |

Note: The sample included LTCI beneficiaries or primary family caregivers. Continuous variables are reported as means and standard deviations. Categorical variables are reported as percentages.

The household monthly income distribution was as follows: 22.7% reported less than CNY 5,000; 35.6% reported CNY 5,001–10,000; 24.1% reported CNY 10,001–20,000; 11.5% reported CNY 20,001–30,000; and 6.1% reported more than CNY 30,000.

Regarding disability level, 62.2% of participants were classified as having mild disability, 26.3% as having moderate disability, and 11.5% as having severe disability. The mean number of chronic conditions was 1.7 (SD = 1.3). The most commonly preferred care model was home-based care, followed by community-based care and institutional care. The mean current monthly personal LTCI premium was CNY 120 (SD = 40). The mean overall satisfaction score for current LTCI services was 3.4 out of 5.0 (SD = 0.8).

Participants were randomly assigned to starting bids of CNY 50, CNY 100, or CNY 150, and starting-bid indicators were included in robustness checks.

### Willingness to pay for improved LTCI services

Table 2 summarizes the WTP results. The mean stated maximum monthly WTP for the predefined LTCI service improvement scenario was CNY 300 (SD = 145), and the median WTP was CNY 250. The mean WTP was higher than the current average monthly personal premium of CNY 120, suggesting that participants, on average, stated a higher valuation for the predefined service improvement scenario than their current reported contribution level.The distribution of monthly WTP is shown in Fig 1.

**Table 2.** Summary of willingness-to-pay findings.

| 表格 |  |  |
| --- | --- | --- |
| Metric | Value | Interpretation |
| Current average personal premium | CNY 120/month | Baseline reported contribution |
| Mean stated WTP for improved services | CNY 300/month | Stated valuation under improvement scenario |
| Median stated WTP | CNY 250/month | Central tendency less affected by right skew |
| 95th percentile of WTP | CNY 800/month | Upper-tail valuation |
| Positive WTP | 96.0% (n = 267) | Participants reporting WTP above zero |
| Genuine zero WTP | 4.0% (n = 11) | Retained as valid zero responses |

Note: The final analytical sample included 278 LTCI beneficiaries or primary family caregivers. No protest-zero responses were included in the final analytical sample.

Among the 278 participants in the final analytical sample, 267 participants reported positive WTP, while 11 participants reported genuine zero WTP. Genuine zero WTP responses were retained in the analysis as left-censored observations. No protest-zero responses were included in the final analytical sample.

The WTP distribution was moderately right-skewed. The median WTP was lower than the mean WTP, and the 95th percentile exceeded CNY 800, indicating that a subset of participants reported relatively high valuations for the described service improvements.

### Factors associated with WTP

Table 3 reports the Tobit regression results. Three main associations were observed.

**Table 3.** Tobit regression results for factors associated with willingness to pay.

| 表格 |  |  |  |
| --- | --- | --- | --- |
| Variable | Coefficient | Robust SE | p value |
| Household monthly income CNY 5,001 – 10,000 | 38.20 | 16.75 | 0.022 |

| Variable | Coefficient | Robust SE | p value |
| --- | --- | --- | --- |
| Household monthly income CNY 10,001 – 20,000 | 82.55 | 18.33 | <0.001 |
| Household monthly income CNY 20,001 – 30,000 | 125.70 | 22.10 | <0.001 |
| Household monthly income > CNY 30,000 | 178.90 | 25.85 | <0.001 |
| Overall satisfaction score, 1 – 5 | –22.15 | 6.88 | 0.001 |
| Age 61 – 75 years | 15.25 | 12.50 | 0.223 |
| Age ≥76 years | 10.18 | 14.22 | 0.474 |
| Moderate disability | 45.33 | 17.89 | 0.011 |
| Severe disability | 68.90 | 19.05 | <0.001 |
| Constant | 152.40 | 28.67 | <0.001 |

Model statistics: Log-likelihood = -1,250.42; sigma = 85.60; left-censored observations = 11; uncensored observations = 267.
Note: Dependent variable: stated monthly WTP in CNY. The reference categories were household monthly income below CNY 5,000, age ≤ 60 years, and mild disability. Additional controls included gender, educational level, urban residence, number of chronic conditions, care preference, policy awareness, respondent type, survey mode, and district fixed effects.

First, household income was positively associated with WTP. Compared with participants with monthly household income below CNY 5,000, participants in higher income groups reported progressively greater WTP. The estimated additional WTP was CNY 38.2 for the CNY 5,001–10,000 group, CNY 82.6 for the CNY 10,001–20,000 group, CNY 125.7 for the CNY 20,001–30,000 group, and CNY 178.9 for the group above CNY 30,000. This pattern is consistent with the possibility that ability to pay may be relevant when considering differentiated contribution arrangements.

Second, satisfaction with current LTCI services was negatively associated with WTP. A one-unit increase in the overall satisfaction score was associated with a decrease of approximately CNY 22.2 in stated WTP, holding other variables constant. One possible interpretation is that participants who were more satisfied with current services perceived less need for additional service improvements.

Third, disability severity was positively associated with WTP. Compared with participants with mild disability, those with moderate disability reported an additional WTP of CNY 45.3, and those with severe disability reported an additional WTP of CNY 68.9. Age group was not statistically significantly associated with WTP in the fully adjusted model.

### Robustness checks

The robustness checks supported the main findings. Ordinary least squares regression using observed WTP produced coefficient estimates that were qualitatively consistent with the Tobit results. Models using log-transformed WTP reduced the influence of upper-tail values but preserved the positive association between income and WTP. Reestimating the Tobit model after excluding genuine zero WTP observations modestly changed the coefficient magnitudes but did not alter the direction or statistical significance of the main predictors. Models including starting-bid indicators did not materially change the main results, suggesting that the randomized starting bids were not driving the observed associations.

## Discussion

### Principal findings

This study estimated WTP for improved LTCI services among LTCI beneficiaries or primary family caregivers in Panjin, a Chinese LTCI pilot city. Three main findings were observed.

First, the mean stated WTP for the predefined LTCI service improvement scenario was approximately CNY 300 per month, compared with the current average monthly premium of approximately CNY 120. This suggests that many participants placed additional value on improvements in LTCI service quality and coverage. However, this result should be interpreted as stated preference under a hypothetical scenario rather than as evidence of actual payment behaviour.

Second, household income was positively associated with WTP. Participants in higher income categories generally reported higher WTP than those in the lowest income group. This finding is consistent with the expectation that ability to pay influences stated demand for health and social insurance improvements [17,18]. It also suggests that uniform contribution increases may affect households differently depending on income level.

Third, disability severity and satisfaction with current services were associated with WTP. Participants with moderate or severe disability reported higher WTP than those with mild disability, suggesting that individuals with greater care needs may value service improvements more. In contrast, higher satisfaction with current LTCI services was associated with lower WTP for additional improvements. One possible interpretation is that participants who were less satisfied perceived greater potential benefit from service upgrades.

### Interpretation and policy relevance

The findings may be relevant to ongoing discussions on LTCI financing in China, where pilot programmes vary in coverage, financing mechanisms, and service delivery arrangements [7,10]. The positive association between income and WTP suggests that differentiated contribution arrangements could be considered in future evaluations of LTCI financing options. However, the study does not identify an optimal premium level, and the results should not be interpreted as evidence that premiums can be increased without affordability concerns.

Any consideration of higher individual contributions would need to account for equity, household financial burden, and the quality of services delivered. The association between lower satisfaction and higher WTP indicates that service quality remains important for public acceptability. Contribution reform without visible improvements in waiting times, caregiver training, care availability, or facility conditions may not be acceptable to beneficiaries or family caregivers.

International experiences, including Japan’s LTCI system, suggest that long-term care financing often requires a combination of public funding, individual contributions, standardized eligibility assessment, and mechanisms for periodic adjustment [11–14,24]. These experiences may offer useful reference points for China, but direct policy transfer should be approached cautiously because institutional arrangements, demographic structures, and fiscal capacities differ across countries.

### Comparison with previous studies

The estimated mean WTP of CNY 300 per month is broadly consistent with stated-preference evidence showing that willingness to contribute to health or long-term care schemes varies with income, perceived need, and expected service quality [18,19]. Previous studies of long-term care financing have emphasized the importance of balancing financial sustainability with affordability and equity [2,10,11,23,24]. Our findings add beneficiary-level evidence from a local Chinese pilot setting and highlight the role of income, disability severity, and satisfaction in shaping stated WTP.

Compared with studies focusing on macro-level financing design or institutional evaluation, this study provides micro-level evidence on how current LTCI beneficiaries or primary family caregivers value potential service improvements. Nevertheless, because the study was conducted in one city and used a stated-preference design, the estimates should be interpreted as context-specific rather than nationally representative.

### Limitations

This study has several limitations. First, the study was conducted in a single LTCI pilot city in northeastern China. Panjin has specific demographic, economic, and policy characteristics, and the findings may not generalize to other regions. Second, WTP was measured using a contingent valuation method and may be affected by hypothetical bias, anchoring effects, strategic responses, or social desirability bias, despite the use of randomized starting bids and a cheap-talk script [19]. Third, the cross-sectional design limits causal interpretation. The observed associations between income, disability severity, satisfaction, and WTP should not be interpreted as causal effects. Fourth, the sample size limited the precision of subgroup analyses. Fifth, distinguishing genuine zero WTP from protest responses may involve some misclassification, depending on participants’ stated reasons for zero WTP [20].

Future studies could use multicity samples, longitudinal designs, discrete choice experiments, or administrative data linkage to examine whether stated WTP corresponds to actual contribution behaviour and service utilization patterns.

## Conclusions

This contingent valuation study found that LTCI beneficiaries or primary family caregivers in Panjin City reported a mean monthly WTP of approximately CNY 300 for a predefined LTCI service improvement scenario, compared with the current average monthly premium of approximately CNY 120. Higher household income and greater disability severity were associated with higher WTP, whereas greater satisfaction with current LTCI services was associated with lower WTP.

These findings suggest that preferences and payment capacity may vary across socioeconomic and care-need groups. The results may inform discussions on differentiated contribution arrangements and service quality improvements in China’s LTCI pilot programmes. However, because the study was based on stated preferences in a single pilot city, the findings should be interpreted cautiously and validated through larger multicity studies and, where possible, evidence on actual payment behaviour.

## Data Availability

Due to ethical restrictions related to participant confidentiality and the scope of informed consent, the individual-level survey data cannot be made publicly available. De-identified data underlying the findings reported in this article are available upon reasonable request from the corresponding author and with permission from the Institutional Review Board of Shenyang Medical Imaging Digital Technology Innovation Center. Data requests may be directed to.

## Data availability statement

The dataset contains potentially sensitive information on disability status, chronic conditions, household income, insurance participation, and service satisfaction. Under the conditions approved by the ethics committee, individual-level data cannot be made publicly available. De-identified data may be made available for researchers who meet the criteria for access to confidential data, subject to approval by the Institutional Review Board of Shenyang Medical Imaging Digital Technology Innovation Center and compliance with applicable privacy regulations. Data requests may be directed to the corresponding author, Xin-yang Li, at.

## Funding

The authors received no specific funding for this work.

## Competing interests

The authors have declared that no competing interests exist.

## Author contributions

Huai-wen Cao: Conceptualization, Data curation, Investigation, Writing – original draft. Xin-yang Li: Conceptualization, Methodology, Supervision, Writing – review & editing. Both authors read and approved the final manuscript.

## Acknowledgments

The authors thank the Panjin Municipal Healthcare Security Bureau for facilitating access to the LTCI beneficiary registry. The authors also thank the community health workers and care facility staff who assisted with participant recruitment and data collection.

---

**Figure 3.**
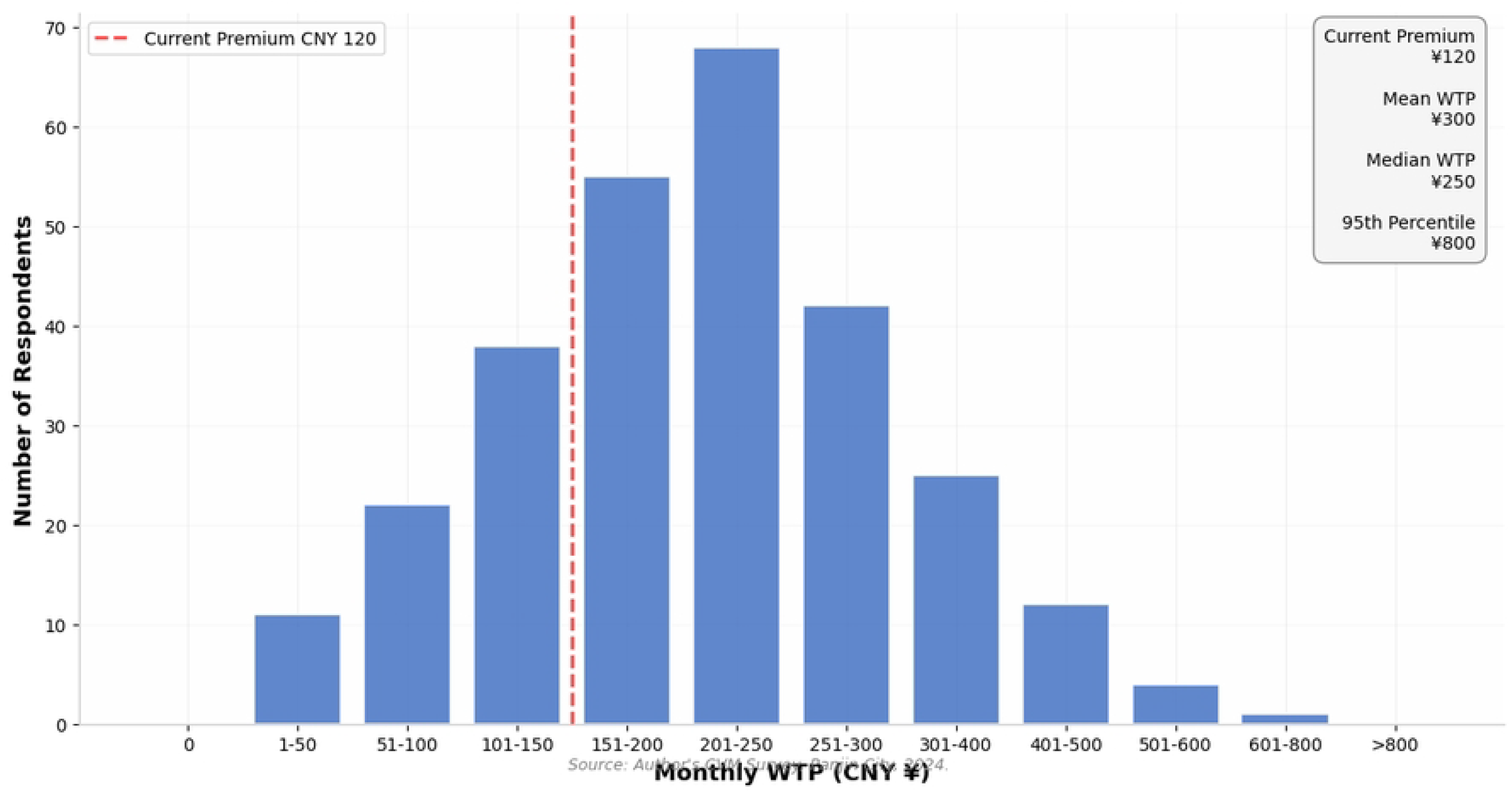
Distribution of monthly willingness-to-pay for improved LTCI services in the Panjin CVM survey (n = 278). Mean WTP = CNY 300, representing 2.5 × the current average premium of CNY 120. The distribution is moderately right-skewed (median = CNY 250, 95th percentile = CNY 800). A total of 96% of respondents expressed positive WTP; 4% stated genuine zero WTP.

## Notes

### Competing Interest Statement

The authors have declared no competing interest.

### Funding Statement

The author(s) received no specific funding for this work.

### Author Declarations

This study was conducted in accordance with the Declaration of Helsinki and was approved by the Institutional Review Board of Shenyang Medical Imaging Digital Technology Innovation Center. Approval number: LNKM20240504-01 All participants provided informed consent prior to participation. The study was conducted in accordance with the principles of the Declaration of Helsinki. The survey data were anonymized before analysis.

